# Challenges With Segmenting Intraoperative Ultrasound For Brain Tumours

**DOI:** 10.1101/2023.12.13.23299820

**Authors:** Alistair Weld, Luke Dixon, Giulio Anichini, Neekhil Patel, Amr Nimer, Michael Dyck, Kevin O’Neill, Adrian Lim, Stamatia Giannarou, Sophie Camp

## Abstract

**Objective:** Addressing the challenges that come with identifying and delineating brain tumours in intraoperative ultrasound. Our goal is to both qualitatively and quantitatively assess the interobserver variation, amongst experienced neuro-oncological intraoperative ultrasound users (neurosurgeons and neuroradiologists), in detecting and segmenting brain tumours on ultrasound. We then propose that, due to the inherent challenges of this task, annotation by localisation of the entire tumour mass with a bounding box could serve as an ancillary solution to segmentation for clinical training, encompassing margin uncertainty and the curation of large datasets.

**Methods:** 30 ultrasound images of brain lesions in 30 patients were annotated by 4 annotators - 1 neuroradiologist and 3 neurosurgeons. The annotation variation of the 3 neurosurgeons was first measured, and then the annotations of each neurosurgeon were individually compared to the neuroradiologist’s, which served as a reference standard as their segmentations were further refined by cross-reference to the preoperative MRI. The following statistical metrics were used: Intersection Over Union, Sørensen-Dice similarity coefficient and Hausdorff distance. These annotations were then converted into bounding boxes for the same evaluation.

**Results:** There was a moderate level of interobserver variance between the neurosurgeons [IoU:0.789, DSC:0.876, HD:103.227] and a larger level of variance when compared against the MRI-informed reference standard annotations by the neuroradiologist, mean across annotators [IoU:0.723, DSC:0.813, HD:115.675]. After converting the segments to bounding boxes, all metrics improve, most significantly, the interquartile range drops by [IoU:37%, DSC:41%, HD:54%].

**Conclusion:** This study highlights the current challenges with detecting and defining tumour boundaries in neuro-oncological intraoperative brain ultra-Sound. We then show that bounding box annotation could serve as a useful complementary approach for both clinical and technical reasons.

## 1 Introduction

Maximal-safe resection of brain tumours is a key pillar of modern neuro-oncology management, improving symptoms, quality of life and overall survival [1]. Accurate delineation of a lesion from surrounding normal functional brain tissue remains challenging but is critical to ensure optimal resection. Intraoperative ultrasound (iUS) is an increasingly popular tool for surgical guidance, as it allows real-time detection, characterisation and outlining of tumours. Furthermore, unlike intraoperative MRI (iMRI), it is easily integrated into the surgical workflow and is relatively affordable. However, adoption, experience and competency with iUS in neurosurgery remain highly variable. There remains a perceived steep learning curve secondary to limited fields of view with unfamiliar topographical representation, artefacts, the unique visuo-tactile task and the difficulty with gaining experience outside of the intraoperative setting. These factors can make learning iUS difficult, with the potential to impair tumour and tumour boundary detection with the risk of leaving unintended residuum or causing inadvertent damage. This challenge is further compounded by the inherent great variation in types of brain lesions, their appearances, the degree of infiltration and the intra-operative changes (such as oedema and haemorrhage) which can further confound. To improve the utility of iUS in neurosurgery, the understanding of the limitations of the tumour-margins delineation capabilities needs to be cemented, and standardised training, new supporting techniques and tools need to be developed.

There are three aims of this study. Firstly, to assess whether tumour boundary detection on iUS is challenging, we will measure interobserver variation between regular iUS operators in segmenting US images of brain lesions. Secondly, we will model the pixel intensities of the segmented tumour boundaries to mathematically model the clarity and blurriness of tumour boundaries. Finally, we will compare this with the interobserver variation of bounding boxes to assess whether this has a role as an alternative, complimentary simplified method for outlining lesion margins. To examine this, we will see whether these bounding boxes can be used as a guide to improve the accuracy of the segmentation.

## 2 Materials and Methods

A preliminary study was conducted using 4 annotators experienced with iUS - a neuroradiologist and 3 neurosurgeons - to determine the foundation of our hypothesis. The neuroradiologist and the three neurosurgeons are all post-training doctors. All clinicians involved have extensive research backgrounds and familiarity with segmentation tools and protocols. Specifically: Neuroradiologist - 10 years of US and neuroimaging experience; An1 - 10 years of neuro-oncology experience with 10 years of *≈* 2-3 cases per week using iUS; An2 - 8 years of neuro-oncology experience with 6 years of *≈* 1 case per week using iUS; An3 - 9 years of neuro-oncology experience with 9 years of *≈* 2 cases per week using iUS. The order of the annotators (An#) has been randomised.

### 2.1 Data

The dataset consists of 30 images, from 30 patients, taken during brain surgery at Charing Cross Hospital. The images were retrospectively selected by the neuroradiologist from cine clips of US sweeps that captured the entire tumour. Images with a field of view which covered the boundaries of the tumour and at least 2cm of surrounding normal brain were selected with reference to the preoperative MRI to ensure accuracy. For patient information please see Tab. 1. The images were acquired using a Canon i900 US machine (Canon Medical Systems, Japan) with a convex probe, and were stored using the Digital Imaging and Communications in Medicine (DICOM) format. All images are of size 960 *×* 1280.

**Table 1:** Patient information. KPS = Karnofsky Performance Status, ASA = American Society of Anesthesiologists Grade, SCC = Squamous Cell Carcinoma, SS = Supratentorial Subependymoma.

| ID | G | Age | Lesion | Location | Histology | KPS | ASA |
| --- | --- | --- | --- | --- | --- | --- | --- |
| 1 | F | 60-69 | Metastasis | Rt Parietal | SCC - lung | 80 | 3 |
| 2 | M | 30-39 | Glioma | Rt Frontal | Astrocytoma | 90 | 1 |
| 3 | M | 50-59 | Glioma | Rt Parietal | Glioblastoma | 80 | 2 |
| 4 | F | 40-49 | Glioma | Lt Frontal, Parietal | Astrocytoma | 90 | 2 |
| 5 | M | 20-29 | Glioma | Lt Parietal | Astrocytoma | 80 | 2 |
| 6 | M | 60-69 | Glioma | Lt Parietal | Glioblastoma | 70 | 2 |
| 7 | M | 50-59 | Glioma | Lt Frontal | Oligodendroglioma | 90 | 3 |
| 8 | M | 50-59 | Glioma | Rt Frontal | Glioblastoma | 90 | 2 |
| 9 | M | 60-69 | Glioma | Lt Parietal | Glioblastoma | 90 | 3 |
| 10 | M | 50-59 | Glioma | Rt Frontal | Glioblastoma | 90 | 2 |
| 11 | M | 40-49 | Glioma | Lt Parietal | Glioblastoma | 80 | 2 |
| 12 | M | 60-69 | Glioma | Rt Parietal | Glioblastoma | 70 | 3E |
| 13 | M | 70-79 | Glioma | Rt Frontal | Glioblastoma | 70 | 2 |
| 14 | M | 10-19 | Granuloma | Lt Temporal | Granuloma TB | 100 | 1 |
| 15 | F | 60-69 | Glioma | Rt Parietal | Glioblastoma | 70 | 2 |
| 16 | M | 60-69 | Glioma | Rt Parietal | Glioblastoma | 80 | 2 |
| 17 | M | 40-49 | Glioma | Lt Temporal | Glioblastoma | 100 | 2 |
| 18 | F | 60-69 | Glioma | Lt Temporal | Glioblastoma | 70 | 2 |
| 19 | M | 40-49 | Glioma | Lt Frontal | Glioblastoma | 80 | 2 |
| 20 | F | 50-59 | Ependymal | Lt Temporal | SS | 90 | 2 |
| 21 | M | 30-39 | Glioma | Post Fossa | Pilocytic astrocytoma | 90 | 1 |
| 22 | M | 60-69 | Glioma | Rt Parietal, Occipital | Glioblastoma | 70 | 2 |
| 23 | F | 60-69 | Glioma | Lt Frontal | Oligodendroglioma | 90 | 2 |
| 24 | M | 30-39 | Glioma | Lt Parietal | Glioblastoma | 90 | 1 |
| 25 | M | 70-79 | Glioma | Rt Parietal | Oligodendroglioma | 90 | 2 |
| 26 | F | 20-29 | Glioma | Rt Frontal | Astrocytoma | 90 | 2 |
| 27 | M | 40-49 | Glioma | Rt Frontal | Astrocytoma | 90 | 2 |
| 28 | F | 10-19 | Glioneuronal | Lt Frontal | DNET | 90 | 1 |
| 29 | F | 40-49 | Glioma | Lt Temporal, Occipital | Oligodendroglioma | 90 | 2 |
| 30 | F | 30-39 | Glioma | Lt Parietal | Astrocytoma | 90 | 2 |

The study had full local ethical approval by the HRA and Health and Care Research Wales (HCRW) authorities. Study title - US-CNS: Multiparametric Advanced Ultrasound Imaging of the Central Nervous System Intraoperatively and Through Gaps in the Bone, IRAS project ID: 275556, Protocol number: 22CX7609, REC reference: 22/WA/0259, Sponsor: Research Governance and Integrity Team (RGIT).

### 2.2 Annotation protocol

The boundaries of tumours were annotated, for both segmentations and bounding boxes, using 3D Slicer (5.4.0) [2]. The annotations provided by the neuroradiologist were made using the benefit of the full US dataset, cross-registration with the preoperative MRI and patient metadata; and we define these as the reference standard/ground truth for this study. The other annotators were given only the individual 2D US images – these annotations were used to evaluate the consistency and accuracy of the annotation capability, comparing inter-neurosurgeon variation and dissimilarity against the annotations of the neuroradiologist. Whilst this is unlike normal clinical practice (where the MRI and full real-time US would be available and employed by the operator) the aim of this study is to assess the ability of B-mode US to delineate brain lesion margins in isolation. The utility of bounding boxes as a guide to refine tumour boundary segmentation was then assessed by An1 repeating their segmentations 3 months later (to mitigate bias) with the reference standard bounding boxes produced by the neuroradiologist overlaid. The bounding box annotations are naively created by taking the maximum and minimum, x and y coordinates from the segments - as opposed to re-annotating.

### 2.3 Statistical analysis

Three statistical metrics were used to compare the similarity between annotations. Within the mathematical framework, let *A* and *B* represent two annotations from the same image i.e. *A* could be the neuroradiologist’s annotation and *B*, one of the neurosurgeon’s annotations. The first two metrics that we define are methods to quantify the degree of overlap - indirectly the shape and volume - between *A* and *B*. The intersection over union (*IoU*) [3] 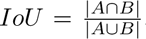, which is defined as the set of image coordinates occupied in the intersection of the two masks, divided by the union of the two masks, the total set of image coordinates occupied by both masks. The Sørensen-Dice similarity coefficient (*DSC*) [4] 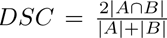, which is defined as twice the intersection of the two masks divided by the cardinalities of the two masks. For both IoU and DSC, the closer the value to 1 the better the score.

For evaluation of the uncertainty of the boundary delineation, the contours of the masks were compared using the Hausdorff distance (HD) [5] 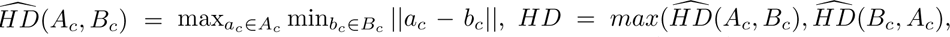 [which determines the maximum Euclidean distance (*pixels*) between all closest point pairs between the two contours sets - the subscript *c* denotes the contours of the corresponding masks, where the contours are the lines intersecting the endpoints of the segment. For this metric, the closer the value is to 0 the better. This analysis was performed using Python, pynrrd [6] for reading the segmentation files and scikit-image [7] and SciPy [8] for the quantitative analysis.

### 2.4 Tumour Border Pixel Dispersion Substudy

To assess the visible distinguishability of the tumour from the surrounding normal tissue, the pixel intensities around the neuroradiologist’s tumour margins are evaluated. By measuring the pixels along the segment’s contour plus a 10 pixels border, perpendicular to, and on both sides, of the contour. To calculate the properties of the distributions, a local maxima, peak-finding method is implemented. Where one peak = unimodal, two = bimodal and three or more = other (could be either multimodal or uniform).

## 3 Results

### 3.1 Inter-Neurosurgeon annotation variance

All results are presented in the following format - Metric[Mean:IQR]. The similarity measure between the neurosurgeon’s annotations is as follows - IoU[0.789:0.115], DSC[0.876:0.072], HD[103.227:73.071]. Although the overlap scores indicate general similarity between the annotations, there are still inconsistencies, which when considering the precision required for tumour resection, this marginal difference can be considered impactful. The average Hausdorff distance, on the other hand, is a significant result. Showing that there is frequent disagreement on at least one point along the tumour boundary.

### 3.2 Tumour Border Pixel Dispersion Substudy

First, we provide a simplified example to illustrate how smoothing affects pixel distributions along a binary boundary (perfect separability). In Fig. 1 a binary circle is created and both Gaussian and box filters are applied with increasing amplification. For the case of the binary circle without smoothing, the boundary is perfectly defined and as such the pixel distribution is bimodal. However, as the smoothing factor increases, the distribution tends towards uniformity or multimodal and under severe blurring, becomes unimodal.

**Figure 1:**
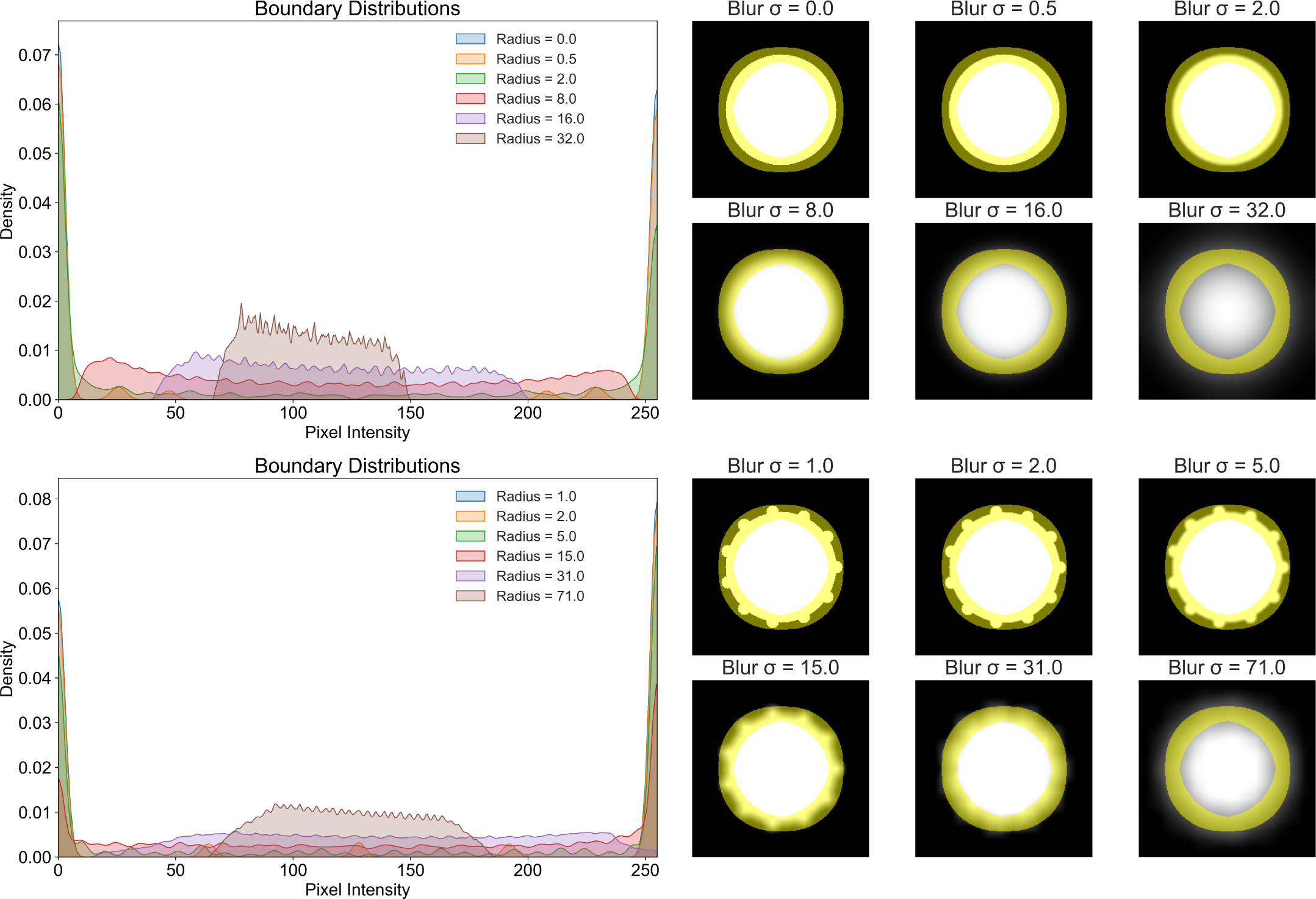
The top row/example shows the effect on the boundary of a binary circle when Gaussian blurring is applied - using a KDE plot. The bottom row/example shows the same but using a box filter, where the circle also now has protrusions, unaffecting the defined boundary represented by the yellow mask.

From evaluating the tumour boundaries, from the 30 images, 28 were classified as unimodal and 2 as bimodal Fig. 2. This result is strong evidence of the severity of the pixel intensity variation along the tumour boundary, which provides mathematical evidence for the ambiguity in defining the discrete, definite boundary points.

**Figure 2:**
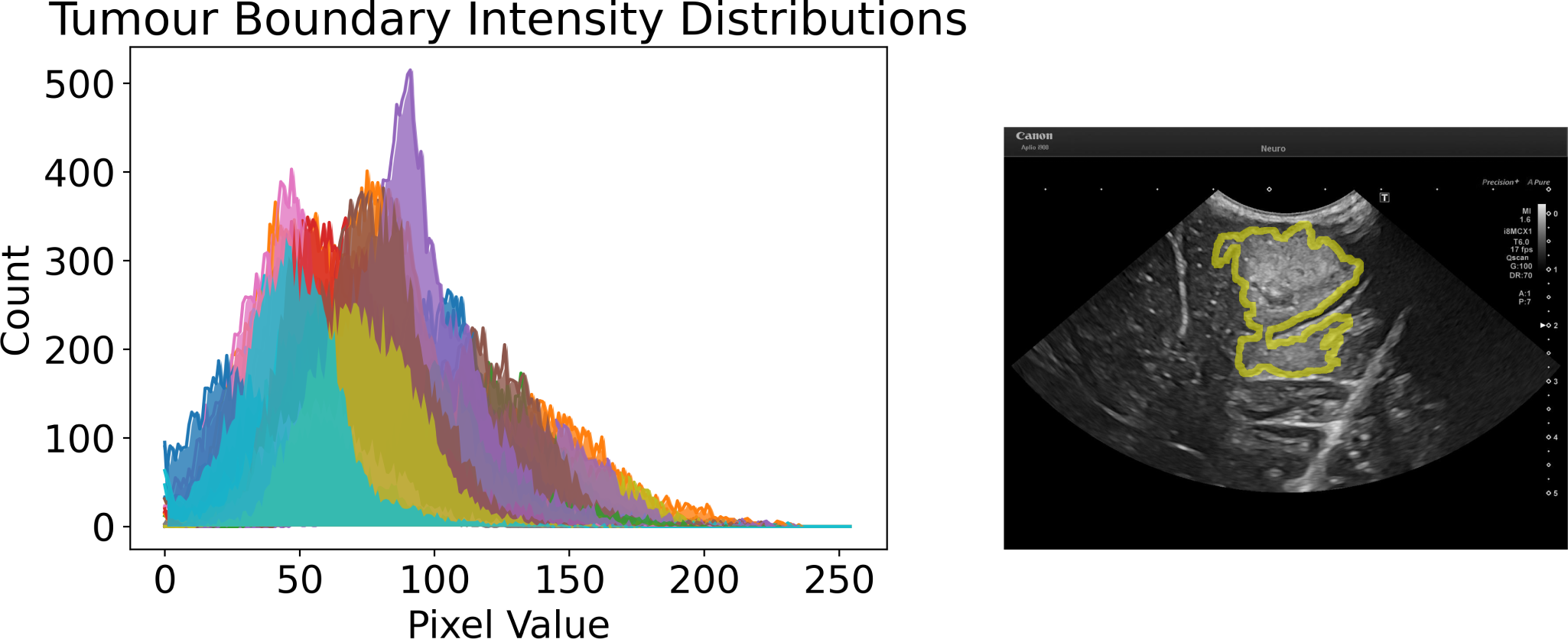
Left is a plot of all tumour boundary pixel intensities distributions. Right is an example of tumour boundary is shown using ID 009.

### 3.3 Neuroradiologist-Neurosurgeon annotation variance

The results of the similarity comparison between the reference standard annotations produced by the neuroradiologist (which benefited from correlation with the preoperative MRI and the full US dataset) and the neurosurgeons is tabulated in Tab. 2,3, and visualised using a box and whiskers plot in Fig. 3. This showed a moderate interobserver variance between the reference standard segmentations and the annotations performed by the neurosurgeon on the single slice B-mode images alone, highlighting the potential limitations and uncertainties of isolated B-mode in defining tumour boundaries.

**Figure 3:**
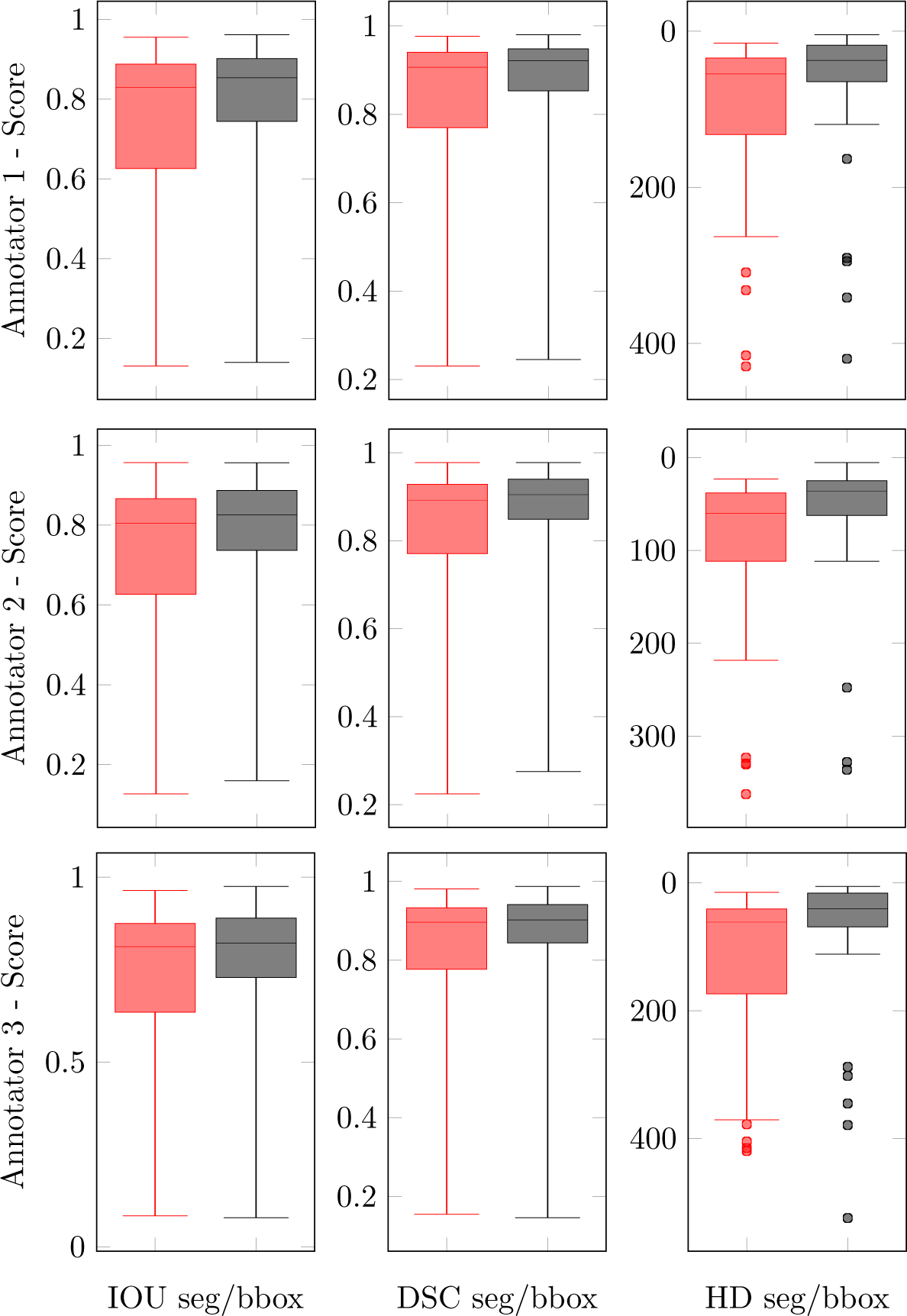
Box and Whisker plots of the annotators IoU, DSC and HD scores on the 30 images - red = segmentation, black = bounding box. What is highlighted is the greatly reduced median score and IQR when using the bounding box method.

**Table 2:** Shown are the IoU and DSC similarity results. The closer the value to 1 the more similar the annotations are.

| Image | An1 |  |  |  | An2 |  |  |  | An3 |  |  |  |
| --- | --- | --- | --- | --- | --- | --- | --- | --- | --- | --- | --- | --- |
|  | Seg |  | BBox |  | Seg |  | BBox |  | Seg |  | BBox |  |
|  | IoU | DSC | IoU | DSC | IoU | DSC | IoU | DSC | IoU | DSC | IoU | DSC |
| 1 | 0.952 | 0.976 | 0.954 | 0.976 | 0.875 | 0.933 | 0.862 | 0.926 | 0.941 | 0.970 | 0.971 | 0.985 |
| 2 | 0.908 | 0.952 | 0.962 | 0.981 | 0.911 | 0.954 | 0.957 | 0.978 | 0.880 | 0.936 | 0.847 | 0.917 |
| 3 | 0.888 | 0.941 | 0.907 | 0.951 | 0.906 | 0.951 | 0.914 | 0.955 | 0.891 | 0.943 | 0.955 | 0.977 |
| 4 | 0.902 | 0.948 | 0.926 | 0.962 | 0.848 | 0.918 | 0.886 | 0.940 | 0.389 | 0.560 | 0.230 | 0.374 |
| 5 | 0.905 | 0.950 | 0.940 | 0.969 | 0.856 | 0.923 | 0.912 | 0.954 | 0.893 | 0.943 | 0.856 | 0.922 |
| 6 | 0.570 | 0.726 | 0.536 | 0.698 | 0.811 | 0.895 | 0.854 | 0.921 | 0.876 | 0.934 | 0.893 | 0.943 |
| 7 | 0.578 | 0.733 | 0.734 | 0.846 | 0.781 | 0.877 | 0.916 | 0.956 | 0.819 | 0.901 | 0.940 | 0.969 |
| 8 | 0.889 | 0.941 | 0.863 | 0.927 | 0.957 | 0.978 | 0.954 | 0.977 | 0.965 | 0.982 | 0.971 | 0.985 |
| 9 | 0.591 | 0.743 | 0.830 | 0.907 | 0.693 | 0.819 | 0.746 | 0.854 | 0.679 | 0.809 | 0.724 | 0.840 |
| 10 | 0.902 | 0.948 | 0.960 | 0.980 | 0.788 | 0.881 | 0.733 | 0.846 | 0.913 | 0.955 | 0.943 | 0.971 |
| 11 | 0.644 | 0.783 | 0.581 | 0.735 | 0.800 | 0.889 | 0.919 | 0.958 | 0.670 | 0.803 | 0.777 | 0.875 |
| 12 | 0.833 | 0.909 | 0.776 | 0.874 | 0.872 | 0.932 | 0.887 | 0.940 | 0.923 | 0.960 | 0.976 | 0.988 |
| 13 | 0.823 | 0.903 | 0.816 | 0.899 | 0.809 | 0.895 | 0.843 | 0.915 | 0.865 | 0.928 | 0.870 | 0.931 |
| 14 | 0.265 | 0.419 | 0.217 | 0.357 | 0.219 | 0.359 | 0.199 | 0.332 | 0.206 | 0.342 | 0.199 | 0.333 |
| 15 | 0.130 | 0.230 | 0.148 | 0.257 | 0.125 | 0.221 | 0.159 | 0.275 | 0.129 | 0.228 | 0.143 | 0.251 |
| 16 | 0.885 | 0.939 | 0.903 | 0.949 | 0.809 | 0.894 | 0.855 | 0.922 | 0.802 | 0.890 | 0.763 | 0.865 |
| 17 | 0.706 | 0.828 | 0.702 | 0.825 | 0.548 | 0.708 | 0.649 | 0.787 | 0.616 | 0.762 | 0.780 | 0.876 |
| 18 | 0.168 | 0.287 | 0.139 | 0.245 | 0.167 | 0.286 | 0.172 | 0.293 | 0.083 | 0.154 | 0.078 | 0.145 |
| 19 | 0.901 | 0.948 | 0.875 | 0.933 | 0.790 | 0.883 | 0.805 | 0.892 | 0.835 | 0.910 | 0.616 | 0.762 |
| 20 | 0.956 | 0.977 | 0.943 | 0.971 | 0.870 | 0.930 | 0.819 | 0.901 | 0.839 | 0.912 | 0.747 | 0.855 |
| 21 | 0.873 | 0.932 | 0.902 | 0.948 | 0.452 | 0.623 | 0.475 | 0.644 | 0.853 | 0.920 | 0.881 | 0.937 |
| 22 | 0.849 | 0.918 | 0.850 | 0.919 | 0.854 | 0.921 | 0.888 | 0.940 | 0.876 | 0.934 | 0.877 | 0.934 |
| 23 | 0.402 | 0.573 | 0.208 | 0.345 | 0.624 | 0.769 | 0.746 | 0.854 | 0.534 | 0.696 | 0.233 | 0.378 |
| 24 | 0.743 | 0.853 | 0.785 | 0.880 | 0.632 | 0.775 | 0.723 | 0.839 | 0.694 | 0.820 | 0.740 | 0.850 |
| 25 | 0.838 | 0.912 | 0.883 | 0.938 | 0.904 | 0.950 | 0.919 | 0.958 | 0.751 | 0.858 | 0.744 | 0.853 |
| 26 | 0.619 | 0.765 | 0.773 | 0.872 | 0.561 | 0.718 | 0.698 | 0.822 | 0.572 | 0.728 | 0.707 | 0.829 |
| 27 | 0.873 | 0.932 | 0.857 | 0.923 | 0.813 | 0.897 | 0.764 | 0.866 | 0.804 | 0.891 | 0.836 | 0.911 |
| 28 | 0.803 | 0.891 | 0.874 | 0.933 | 0.773 | 0.872 | 0.832 | 0.908 | 0.756 | 0.861 | 0.908 | 0.952 |
| 29 | 0.791 | 0.883 | 0.867 | 0.929 | 0.620 | 0.766 | 0.754 | 0.859 | 0.622 | 0.767 | 0.807 | 0.893 |
| 30 | 0.821 | 0.902 | 0.810 | 0.895 | 0.878 | 0.935 | 0.816 | 0.899 | 0.832 | 0.908 | 0.879 | 0.936 |
| Mean | 0.734 | 0.821 | 0.751 | 0.827 | 0.719 | 0.812 | 0.755 | 0.837 | 0.717 | 0.807 | 0.730 | 0.808 |

**Table 3:** Shown are the HD similarity results. The closer the value to 0 pixels the better aligned the annotation margins are.

| Image | An1 |  | An2 |  | An3 |  |
| --- | --- | --- | --- | --- | --- | --- |
|  | Seg | BBox | Seg | BBox | Seg | BBox |
| 1 | 14.765 | 8.544 | 43.000 | 36.620 | 14.142 | 5.000 |
| 2 | 22.204 | 4.123 | 25.000 | 5.000 | 52.802 | 46.271 |
| 3 | 33.734 | 16.492 | 32.558 | 14.142 | 23.324 | 9.220 |
| 4 | 29.069 | 8.246 | 42.438 | 33.061 | 371.389 | 379.120 |
| 5 | 51.614 | 13.153 | 57.245 | 19.416 | 40.162 | 23.345 |
| 6 | 162.926 | 163.515 | 56.223 | 35.511 | 52.924 | 17.804 |
| 7 | 74.632 | 40.311 | 32.650 | 7.280 | 42.579 | 6.000 |
| 8 | 52.000 | 48.010 | 33.121 | 7.616 | 28.636 | 6.403 |
| 9 | 137.773 | 49.477 | 74.686 | 47.265 | 79.202 | 48.836 |
| 10 | 25.807 | 4.243 | 55.803 | 44.598 | 18.682 | 6.403 |
| 11 | 117.652 | 119.620 | 362.627 | 15.000 | 138.105 | 69.231 |
| 12 | 66.000 | 70.000 | 68.447 | 47.170 | 43.600 | 7.071 |
| 13 | 46.174 | 28.844 | 62.000 | 30.000 | 40.522 | 23.324 |
| 14 | 331.724 | 341.264 | 329.524 | 336.265 | 331.965 | 345.307 |
| 15 | 263.610 | 294.703 | 218.563 | 247.746 | 254.342 | 288.043 |
| 16 | 36.770 | 36.000 | 81.006 | 44.000 | 90.554 | 68.710 |
| 17 | 309.015 | 110.000 | 330.510 | 102.000 | 377.922 | 62.169 |
| 18 | 429.439 | 419.640 | 323.303 | 328.056 | 404.901 | 524.675 |
| 19 | 27.295 | 23.000 | 58.052 | 36.401 | 419.715 | 112.058 |
| 20 | 30.887 | 23.000 | 64.938 | 58.694 | 87.727 | 84.000 |
| 21 | 42.012 | 15.000 | 119.549 | 111.973 | 33.377 | 14.866 |
| 22 | 68.447 | 28.425 | 70.264 | 25.000 | 43.600 | 24.000 |
| 23 | 415.473 | 290.493 | 57.079 | 44.721 | 415.120 | 302.154 |
| 24 | 71.007 | 48.703 | 89.022 | 63.789 | 80.000 | 53.488 |
| 25 | 56.511 | 20.809 | 22.627 | 14.213 | 63.600 | 35.468 |
| 26 | 156.259 | 45.188 | 171.047 | 70.767 | 175.026 | 57.271 |
| 27 | 15.000 | 10.440 | 24.000 | 24.413 | 29.547 | 21.024 |
| 28 | 34.670 | 30.067 | 36.056 | 30.017 | 35.228 | 13.601 |
| 29 | 53.254 | 38.949 | 180.049 | 79.649 | 171.234 | 54.406 |
| 30 | 67.231 | 40.792 | 26.926 | 27.731 | 59.641 | 24.739 |
| Mean | 108.098 | 79.702 | 104.94 | 66.270 | 133.986 | 91.134 |

With conversion of the segments to bounding boxes there was a noticeable improvement in variance with a sizable decrease in the interquartile range (IQR). For all annotators, the naive conversion to a bounding box, overall, increases the annotation similarity. The decrease in the IQR, as a percentage, for [IoU, DSC, HD] are as follows - An1=[40%, 44%, 52%], An2=[37%, 42%, 49%], An3=[33%, 37%, 60%].

Further evaluation of the bounding box proposal is conducted by measuring individually for each image, the percentage of the neuroradiologist’s segmentation contained within the neurosurgeon’s corresponding bounding box. The average results per annotator are - An1:98.387%, An2:98.833%, An3:99.052%. From the results, it can be concluded that the bounding box approach is usable for localising the entire tumour mass, and is a suitable method for reducing inter-observer annotation variance.

### 3.4 Qualitative observations

Sample images are displayed in Fig. 4,5. For all images: original image = left, neuroradiologist annotation = middle, neurosurgeons annotation = right. An assortment of different sources of boundary aleatoric uncertainty are shown in Fig. 4. Including images containing fuzzy borders and continued hyperechogenicity extending beyond the tumour. Shown in Fig. 5 are example cases where using bounding boxes has substantially improved the annotation similarity.

**Figure 4:**
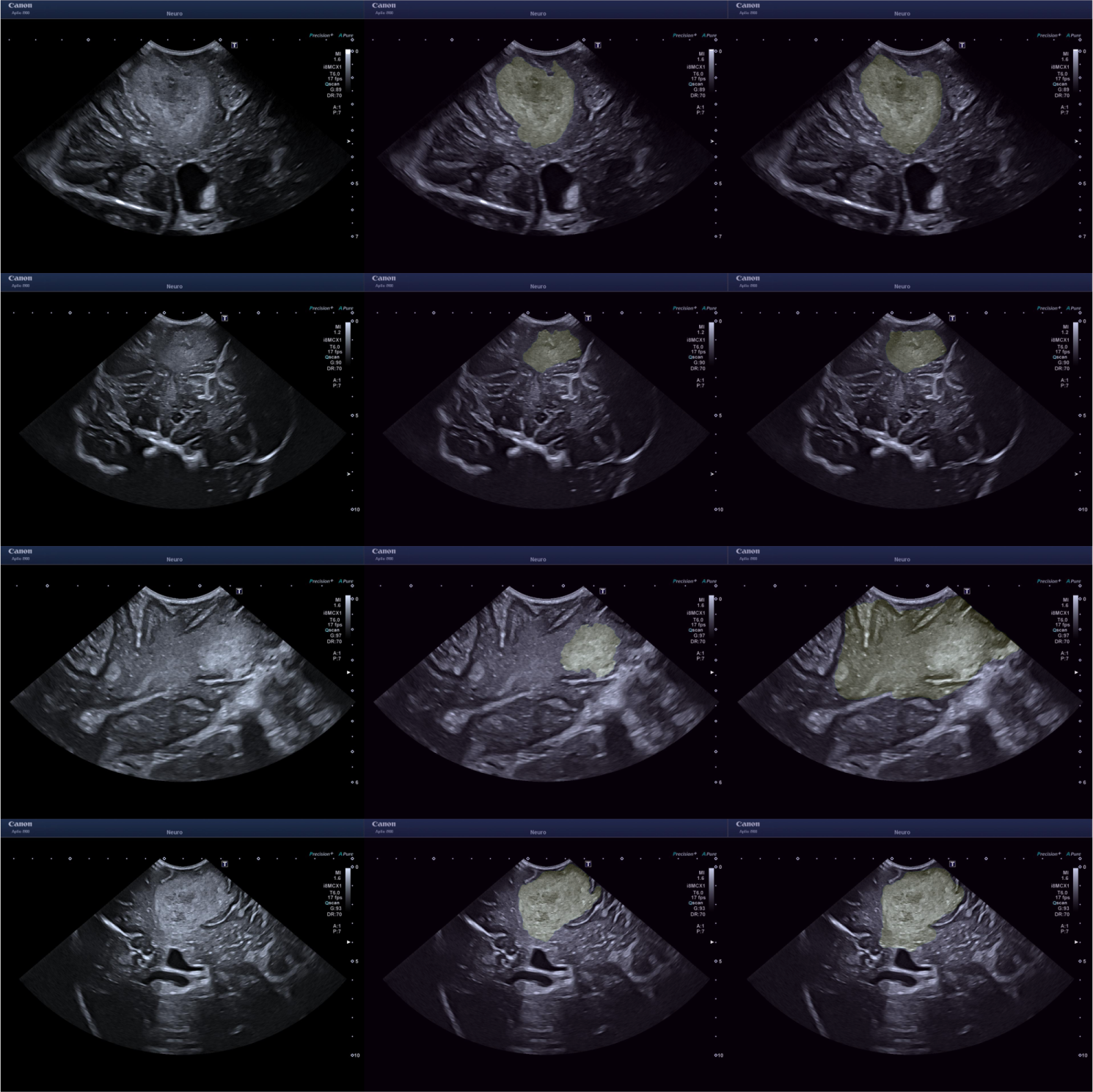
Uncertainty caused by unclear boundary. The left images are the original images, the middle from the neuroradiologist and the right from a neurosurgeon. From top to bottom - ID 13 An3, ID 07 An3, ID 18 An2, ID 19 An2.

**Figure 5:**
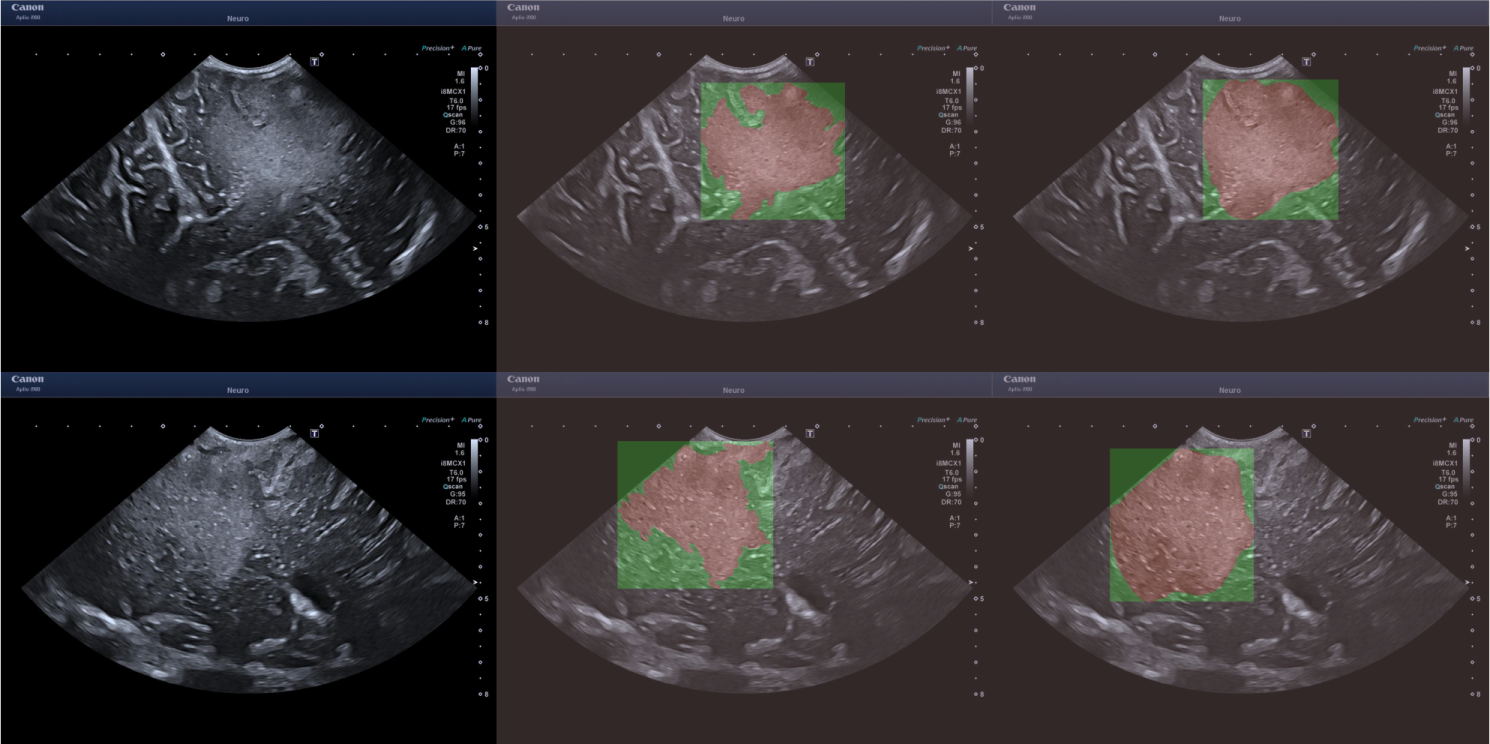
Examples for where the segments have large dissimilarity whilst the bounding boxes don’t. The left images are the original images, the middle from the neuroradiologist and the right from a neurosurgeon. From top to bottom - ID 11 AN 2, the bottom from ID 26 AN 1.

### 3.5 Improving segmentation accuracy using overlaid bounding boxes as a guide

In the substudy looking at the impact of overlaid reference bounding boxes on An1 segmentation accuracy, we found a substantial improvement in segmentation similarity between the neurosurgeon and reference standard. The previous results compared to the new results are as follows - without bounding box [IOU:0.734, DSC:0.821, HD:108.098], with bounding box [IOU:0.858, DSC:0.922, HD:52.072]. This result shows that the bounding box can be used as a visual anchor to minimise the uncertainty when segmenting.

## 4 Discussion

Intraoperative US (iUS) has been employed in neurosurgery for 70 years [9]. Over the last two decades the adoption of iUS has greatly increased, in part due to the advent of modern high-quality US and navigated US co-registered with preoperative MRI and CT [10–14]. Despite these improvements, iUS adoption in brain tumour surgery is not universal [15]. Although iUS is relatively affordable and readily deployed in most theatre settings, there remains a perceived steep learning curve and concerns regarding the accuracy of iUS. Presently, there have been only a few meta-analyses looking at the impact of iUS in glioma surgery. One pooled series reported an average 77% gross total resection rate in 739 patients undergoing iUS-guided resection (71.9% in HGG compared to 78.1% in LGG) which was comparable to other forms of navigation. A recent meta-analysis of 409 diffuse gliomas compared the accuracy of iUS to the reference standard post-operative MRI. They found that iUS was an effective technique in assessing diffuse glioma resection, with a 72.2% pooled sensitivity and a 93.5% pooled specificity [16]. Whilst these results are encouraging the current evidence supports a need to improve iUS accuracy if it is to become part of the standard of care in brain tumour surgery. Trials assessing the role of US in neurosurgery, such as the randomized controlled trial Functional and Ultrasound-Guided Resection of Glioblastoma (FUTURE-GB), and refining of US techniques are therefore needed [17]. In this study, we further highlight the specific challenge of tumour detection and tumour boundary delineation in cranial iUS. Here we demonstrate that there remains moderate to high interobserver variation in the identification and segmentation of tumours on B-mode images acquired on a modern, current-generation US scanner, between four individuals with experience in iUS-guided brain surgery. Three broad elements likely contribute to this variance, 1) the specific qualities of brain tumours, 2) technical ultrasound factors and 3) operator influences.

Firstly, there are inherent features of brain tumours themselves which can make them difficult to delineate. Gliomas, in particular, are well-known to be infiltrative tumours, meaning that they spread cancer cells beyond their obvious radiological margins [18, 19]. Moreover, both high-grade gliomas and low-grade gliomas often show a degree of surrounding oedema which is, to this day, challenging to interpret in terms of differential diagnosis between reactive inflammatory tissue or actually infiltrated brain, even using more established neuroimaging tools such as MRI. One of the main points of discrepancy in the present series was the definition of tumour margins. For example, in some cases, the lack of clarity over whether the highlighted hyperechogenicity of the tissue was caused by a continuation of the tumour mass or just reactive oedema, introduces significant heteroscedastic aleatoric uncertainty when defining the exact margin. This challenge is further emphasised by how there remains no reference standard imaging technique that absolutely defines tumour extent, nor is it usually possible to remove tumours en bloc in intra-axial neurosurgery, precluding accurate histological correlation of tumour margins. Secondly, there are unique challenges that US presents. There are numerous ways that an ultrasound image can be altered, including changes in settings (such as gain and frequency), ultrasound machine, probe type, probe contact and probe angle. In most cases, several of these parameters need to be intentionally tailored to the particular tumour being imaged. For instance, using a low-frequency probe, which has a trade-off in reduced spatial resolution, to visualise a deep tumour versus a high-frequency probe, with high spatial resolution, to image a superficial cortical tumour. This wide range of US options can greatly alter the final image creating another source of variance and in turn, aleatoric uncertainty which is arguably greater than typically seen with other established imaging modalities such as CT and MRI. US is also vulnerable to several unique artefacts, such as acoustic shadowing [20] and acoustic enhancement, which can alter and obscure the image creating a further source of uncertainty. These issues could be mitigated somewhat by the establishment of a standardised protocol for US settings and image acquisition. However, even then, it would be impossible to fully account for all scenarios due to the wide spectrum of tumours and anatomical locations. The potential for confounding artefacts and uncertainty regarding tumour boundaries further increases as surgery progresses due to increased deformation, oedema and potential obscuring blood products. This uncertainty could also be reduced with operator experience, which links to operator factors which are the final source of variance. Currently, neurosurgical training in US is predominantly experiential based on exposure to live cases in theatres. Whilst this is an essential aspect of learning a new surgical skill this can greatly prolong the learning phase due to a relatively low rate of exposure to the imaging technique as it necessitates an intraoperative setting with a craniotomy window. This is in contrast to CT and MRI, which neurosurgeons are much more comfortable with interpreting, owing, in part, to these being readily available and performed regularly on most patients.

There are several complementary ways that iUS accuracy could be improved and the steep learning curve could be flattened and shortened. This includes the use of advanced multimodal US (including contrast-enhanced US) and navigated US. In addition, dedicated courses employing US phantoms with brain tumour models can greatly help, by providing both applied formal training in US theory in addition to hands-on time with US scanning. In all cases, however, these approaches still require significant time investment and training. In this context, automated, computer-assisted, detection and segmentation of brain tumours on ultrasound would be a desirable and useful additional tool. Automated segmentation of brain tumours on MRI has been well explored with several robust and open source tools now available [21]. In contrast, development in automated segmentation of iUS images is in its infancy with no applications yet available. There are several reasons for this. As illustrated in this study it is challenging to establish a ground truth data set using manual segmentation due to high interobserver variance. Furthermore, unlike MRI there is a paucity of neurooncology iUS imaging datasets this is likely due to the relatively low number of iUS scans acquired, the greater logistical challenges with saving and downloading scans, the potentially large file sizes of video acquisitions, the greater variation of iUS across sites and the need for highly experienced annotators to perform the time-consuming segmentations. Considering these many boundaries, we are far off the realisation of a reliable system to rapidly automatically and accurately segment iUS brain tumour images. To bridge this gap, here we assess the utility of bounding boxes as an additional complementary tool for simplified tumour detection and delineation.

Unsurprisingly, we found much lower interobserver variation when using bounding boxes to define tumour location and margins compared to segmentations. The advantage of our proposed use of bounding boxes is a step in the direction of overcoming the above-mentioned challenges. From a clinical perspective, the bounding box would be useful for training purposes and immediate identification of the tumour mass. While this system is expected to be lower in specificity, the high sensitivity should assist inexperienced surgeons in detecting tumours and providing an area to focus on. Further, the whole signal change (fuzzy margin) would be included in the bounding box, thus making sure that there is no missing tumour from the targeted area. The process of annotation is also much quicker for bounding boxes although this will be annotator-dependent, from our experience, bounding box annotation may take as little as 1/3 of the time of segmentation, reducing the manual labour cost.

In computer vision and AI, segmentation methods will typically define/assume that the segmentation task can be framed as separating an image into sub-regions with defined and complete boundaries. For example, explicitly through the definition of a mathematical optimisation framework, or implicitly by training a neural network on common datasets. Because of this, the task of semantically segmenting brain tumours in US becomes uniquely difficult. From an engineering perspective, there are a large number of benefits to using bounding boxes. First and foremost, the reduced annotation complexity should facilitate the collection of large datasets. Accelerating technical and clinical research into this topic. For technical development, the primary benefit is the reduced complexity of the estimation task, which should lead to highly accurate systems. The black boxes can also be used for different tasks such as: representing the segment as a probabilistic heat map [22] to account for the infiltrativity [18, 19], prompting large/powerful models such as Segment Anything Model (SAM) [23], tracking algorithms [24].

There are a few limitations of this study. Firstly, the number of annotated cases was small which is why we have withheld from identifying correlates - such as whether certain tumour types are more accurately segmented. Secondly, only single slices were used for segmentation as opposed to volumes. This is unlike the real-world use of iUS where assessment of boundaries is based on live 3D sweeps of tumours and adjacent anatomy, plus often cross-correlation with preoperative MRI, both of which would help refine the accuracy of segmentations. Extending from this, there remains the recurring issue in neuro-oncology of there not being an accepted gold-standard ground truth for tumour boundaries and our use of integrated multimodal MRI and US to create the reference standard segmentation’s has to be an accepted compromise. Finally, whilst bounding boxes may serve as an efficient method to improve the detection of tumours, this is at the expense of specificity, which is important for the prevention of inadvertent removal of normal, functional brain tissue.

## 5 Conclusion

What has been highlighted in this paper is our thoughts on the challenges of segmenting brain tumours in 2D US images, with a preliminary study conducted to corroborate our hypothesis. For future work, larger curated and consensus annotated data sets of iUS brain tumour images and volumes are needed to develop more accurate computer assisted boundary detection tools. This is likely to only be achieved through multi-site collaboration and pooling of data.

## Data Availability

All data produced will not be made available

## Acknowledgements

This research was supported by the UK Research and Innovation (UKRI) Centre for Doctoral Training in AI for Healthcare (EP/S023283/1), the Royal Society (URF*\*R*\*2 01014]), the NIHR Imperial Biomedical Research Centre, Canon Medical Systems and Brain Tumour Research (BTR) charity.

## References

[1] Hervey-Jumper, S.L., Berger, M.S.: Maximizing safe resection of low-and high-grade glioma. Journal of Neuro-Oncology 130, 269–282 (2016)

[2] Fedorov, A., Beichel, R., Kalpathy-Cramer, J., Finet, J., Fillion-Robin, J.-C., Pujol, S., Bauer, C., Jennings, D., Fennessy, F., Sonka, M., Buatti, J., Aylward, S., Miller, J.V., Pieper, S., Kikinis, R.: 3d slicer as an image computing platform for the quantitative imaging network. Magn. Reson. Imaging 30, 1323–1341 (2012)

[3] Rezatofighi, S.H., Tsoi, N., Gwak, J., Sadeghian, A., Reid, I.D., Savarese, S.: Generalized intersection over union: A metric and a loss for bounding box regression. 2019 IEEE/CVF Conference on Computer Vision and Pattern Recognition (CVPR), 658–666 (2019)

[4] Hofmanninger, J., Prayer, F., Pan, J., Röhrich, S., Prosch, H., Langs, G.: Automatic lung segmentation in routine imaging is primarily a data diversity problem, not a methodology problem. European Radiology Experimental 4 (2020)

[5] Karimi, D., Salcudean, S.E.: Reducing the hausdorff distance in medical image segmentation with convolutional neural networks. IEEE Transactions on Medical Imaging 39, 499–513 (2019)

[6] Everts, M., Elliott, A., Braun-Jones, T., Court, R., Johnson, H., Norton, I., Warner, J., Fischer, P., Meine, H., Ghayoor, A., Billah, T., Ekström, S., Leinweber, K., JcNils, Gronde J., Brown, D.: mhe/pynrrd: v0.4.1 released (2019) 10.5281/zenodo.3544486

[7] Walt, S., Schönberger, J.L., Nunez-Iglesias, J., Boulogne, F., Warner, J.D., Yager, N., Gouillart, E., Yu, T., contributors: scikit-image: image processing in Python. PeerJ 2, 453 (2014) 10.7717/peerj.453

[8] Virtanen, P., Gommers, R., Oliphant, T.E., Haberland, M., Reddy, T., Cournapeau, D., Burovski, E., Peterson, P., Weckesser, W., Bright, J., van der Walt, S.J., Brett, M., Wilson, J., Millman, K.J., Mayorov, N., Nelson, A.R.J., Jones, E., Kern, R., Larson, E., Carey, C.J., Polat, I., Feng, Y., Moore, E.W., VanderPlas, J., Laxalde, D., Perktold, J., Cimrman, R., Henriksen, I., Quintero, E.A., Harris, C.R., Archibald, A.M., Ribeiro, A.H., Pedregosa, F., van Mulbregt,P., SciPy 1.0 Contributors: SciPy 1.0: Fundamental Algorithms for Scientific Computing in Python. Nature Methods 17, 261–272 (2020) 10.1038/ s41592-019-0686-2

[9] French, L.A., Wild, J.J., Neal, D.: Detection of cerebral tumors by ultrasonic pulses. pilot studies on postmortem material. Cancer 3(4), 705–708 (1950)

[10] Dohrmann, G.J., Rubin, J.M.: History of intraoperative ultrasound in neurosurgery. Neurosurgery Clinics of North America 12(1), 155–66 (2001)

[11] Hata, N., Dohi, T., Iseki, H., Takakura, K.: Development of a frameless and armless stereotactic neuronavigation system with ultrasonographic registration. Neurosurgery 41(3), 608–13 (1997)

[12] Ji, S., Roberts, D.W., Hartov, A., Paulsen, K.D.: Intraoperative patient registration using volumetric true 3d ultrasound without fiducials. Medical physics 39(12), 7540–7552 (2012)

[13] Ohue, S., Kumon, Y., Nagato, S., Kohno, S., Harada, H., Nakagawa, K., Kikuchi, K., Miki, H., Ohnishi, T.: Evaluation of intraoperative brain shift using an ultrasound-linked navigation system for brain tumor surgery. Neurologia medico-chirurgica 50(4), 291–300 (2010)

[14] Mahboob, S.O., McPhillips, R., Qiu, Z., Jiang, Y., Meggs, C., Schiavone, G., Button, T.W., Desmulliez, M.P.Y., Demore, C., Cochran, S., Eljamel, S.: Intraoperative ultrasound-guided resection of gliomas: A meta-analysis and review of the literature. World neurosurgery 92, 255–263 (2016)

15. Dixon, L., Lim, A., Grech-Sollars, M., Nandi, D., Camp, S.J.: Intraoperative ultrasound in brain tumor surgery: A review and implementation guide. Neurosurgical Review 45, 2503–2515 (2022)

[16] Trevisi, G., Barbone, P., Treglia, G., Mattoli, M.V., Mangiola, A.: Reliability of intraoperative ultrasound in detecting tumor residual after brain diffuse glioma surgery: a systematic review and meta-analysis. Neurosurgical Review 43, 1221– 1233 (2019)

[17] Plaha, P., Camp, S., Cook, J., McCulloch, P., Voets, N., Ma, R., Taphoorn, M.J.B., Dirven, L., Grech-Sollars, M., Watts, C., Bulbeck, H., Jenkinson, M.D., Williams, M., Lim, A., Dixon, L., Price, S.J., Ashkan, K., Apostolopoulos, V., Barber, V.S., Taylor, A., collaborators, F.-G., Nandi, D.: Future-gb: functional and ultrasound-guided resection of glioblastoma – a two-stage randomised control trial. BMJ Open 12(11) (2022) 10.1136/bmjopen-2022-064823 https://bmjopen.bmj.com/content/12/11/e064823.full.pdf

[18] Pallud, J., Audureau, E., Blonski, M., Sanai, N., Bauchet, L., Fontaine, D., Mandonnet, E., Dezamis, E., Psimaras, D., Guyotat, J., et al.: Epileptic seizures in diffuse low-grade gliomas in adults. Brain 137(2), 449–462 (2014)

[19] Sahm, F., Capper, D., Jeibmann, A., Habel, A., Paulus, W., Troost, D., Von Deimling, A.: Addressing diffuse glioma as a systemic brain disease with single-cell analysis. Archives of neurology 69(4), 523–526 (2012)

[20] Weld, A., Dixon, L., Anichini, G., Dyck, M., Ranne, A., Camp, S.J., Giannarou, S.: Identifying visible tissue in intraoperative ultrasound images during brain surgery: A method and application. ArXiv abs/2306.01190 (2023)

[21] Pati, S., Baid, U., Edwards, B., et al.: Federated learning enables big data for rare cancer boundary detection. Nature Communications 13 (2022)

[22] Petsiuk, V., Jain, R., Manjunatha, V., Morariu, V.I., Mehra, A., Ordonez, V., Saenko, K.: Black-box explanation of object detectors via saliency maps. 2021 IEEE/CVF Conference on Computer Vision and Pattern Recognition (CVPR), 11438–11447 (2020)

[23] Kirillov, A., Mintun, E., Ravi, N., Mao, H., Rolland, C., Gustafson, L., Xiao, T., Whitehead, S., Berg, A.C., Lo, W.-Y., Dolár, P., Girshick, R.B.: Segment anything. ArXiv abs/2304.02643 (2023)

[24] Cartucho, J., Weld, A., Tukra, S., Xu, H., Matsuzaki, H., Ishikawa, T., Kwon, M.S., Jang, Y.E., Kim, K.-J., Lee, G., Bai, B., Kahrs, L.A., Boecking, L., Allmendinger, S., Muller, L., Zhang, Y., Jin, Y., Sophia, B., Vasconcelos, F., Reiter, W., Hajek, J., Silva, B.L.B., Buschle, L.R., Lima, E., Vilaça, J.L., Queiós, S., Giannarou, S.: Surgt challenge: Benchmark of soft-tissue trackers for robotic surgery. Medical image analysis 91, 102985 (2023)

